# CT-based imaging metrics for identification of radiation-induced lung damage

**DOI:** 10.1101/2022.12.18.22283636

**Authors:** Joonsang Lee, Marcelo Benveniste, Erika G. Odisio, Laurence E. Court, Steven H. Lin

## Abstract

**Purpose:** This study investigates the feasibility of radiomics for identifying textural changes of radiation-induced lung damage (RILD) after chemoradiotherapy.

**Methods:** The severity of RILD on each CT scan was graded on a scale from 0 (similar to the baseline CT scan) to 5 (lung fibrosis). The delineation of abnormal areas inside the lung on CT images was performed semi-automatically using a median filter. We extracted a total of 138 quantitative image features from this delineated region of interest and ran a random forest algorithm as a classifier for identifying the severity of RILD. After training and testing the model, we validated the model using a separate dataset.

**Results:** The classification accuracies for identifying grade 0 from grades 1 ∼ 5 were 70% for the test dataset and 85% for the validation dataset; for identifying grade 1 from grades 2∼5, 90% for the test dataset and 95% for the validation dataset; and for identifying grade 5 from grades 2∼4, 80% for the test dataset and 85% for the validation dataset.

**Conclusions:** Our preliminary study shows that the classification accuracy was robust, the model was most useful for distinguishing grade 1 from other grades, and the results demonstrated the feasibility of radiomics for identifying the severity of lung damage after chemoradiotherapy. This approach could be a potential tool for helping diagnostic radiologists identify RILD and its severity on CT images.

## INTRODUCTION

The alveolar/capillary complex of the lung is radiosensitive, and patients who receive chemo-radiotherapy for lung cancer may have a risk of lung damage. The occurrence of radiation-induced lung damage (RILD) varies depending on tumor location, type, and treatment modality^1-5^. Five to thirty percent of patients who receive radiotherapy (RT) for thoracic malignancies develop symptomatic radiation pneumonitis, and 50% to 100% of patients have radiologic evidence of regional injury ^6-8^. RILD can be separated into early and late effects of radiation. Early radiation effects manifest as radiation pneumonitis, and late RILD typically presents as pulmonary fibrosis ^*9-11*^.

The severity of lung damage varies widely among patients. RILD can have a variety of appearances on CT images, depending on the severity of damage. Several studies have been conducted to predict and quantify RILD, such as investigating regional lung density changes to assess the dose-dependent nature of these changes for patient- and treatment-associated factors ^*12*^, investigating an individual patient data meta-analysis to determine factors of clinically significant pneumonitis ^*3,13*^, and investigating both the physical and biologic parameters V_30_ to predict the risk of symptomatic radiation-induced lung injury^*14*^. These studies are based on mean lung CT density or biologic parameters. Sets of higher-order measurements using texture analysis have also been used to capture the complexity of lung disease patterns. Several textural studies have demonstrated the utility of texture features to quantify lung disease; the ability to classify lung diseases with centrilobular emphysema, panlobular emphysema, and constrictive obliterative bronchiolitis^15^; and the ability to visualize lung texture in serial thoracic CT scans^16^; and the ability of texture analysis to identify patients who develop radiation pneumonitis^17^.

Human interpretation of medical images is limited and impractical especially when clinicians need to study thousands of medical images. Radiomics is an important revolution in visually identifiable imaging technology and an emerging field in medical study for extracting a large amount of quantitative image features, including intensity features, texture features from co-occurrence matrices or run-length matrix features, and shape features. The purpose of this study was to examine the feasibility of radiomics for identifying CT textural changes in the lung after chemo-radiotherapy. Further, this preliminary study could help to find CT based imaging predictors of RILD by capturing early CT textural changes in the lung after chemo-radiotherapy that could predict for RILD for the future study.

## METHODS

### Patient and imaging data

A total of 195 lung cancer patients at The University of Texas MD Anderson Cancer Center between February 2001 and October 2012 were identified under IRB approval. These patients had diagnostic CT scans taken before and after RT. The slice thickness varied from 1.3mm to 7.5mm; in this study we attempted to minimize noise sources by only using 2.5mm and 3.8mm slice thicknesses. After exclusion on slice thicknesses, a total of 86 lung cancer patients were investigated. We selected CT scans randomly from this set of patients to create a training and test data cohort (110 scans), and a validation cohort (60 scans). CT images were acquired with a 512 × 512 image matrix and multiple voxel sizes of 0.7∼0.98 mm x 0.7∼0.98 mm x 2.5∼3.8 mm and were reconstructed using the lung kernel. All images were resampled to an isotropic voxel size of 1 mm x 1 mm x 2.5 mm using trilinear interpolation before we analyzed the data. Table 1 summarize patients’ characteristics and radiation dose.

**TABLE 1.**
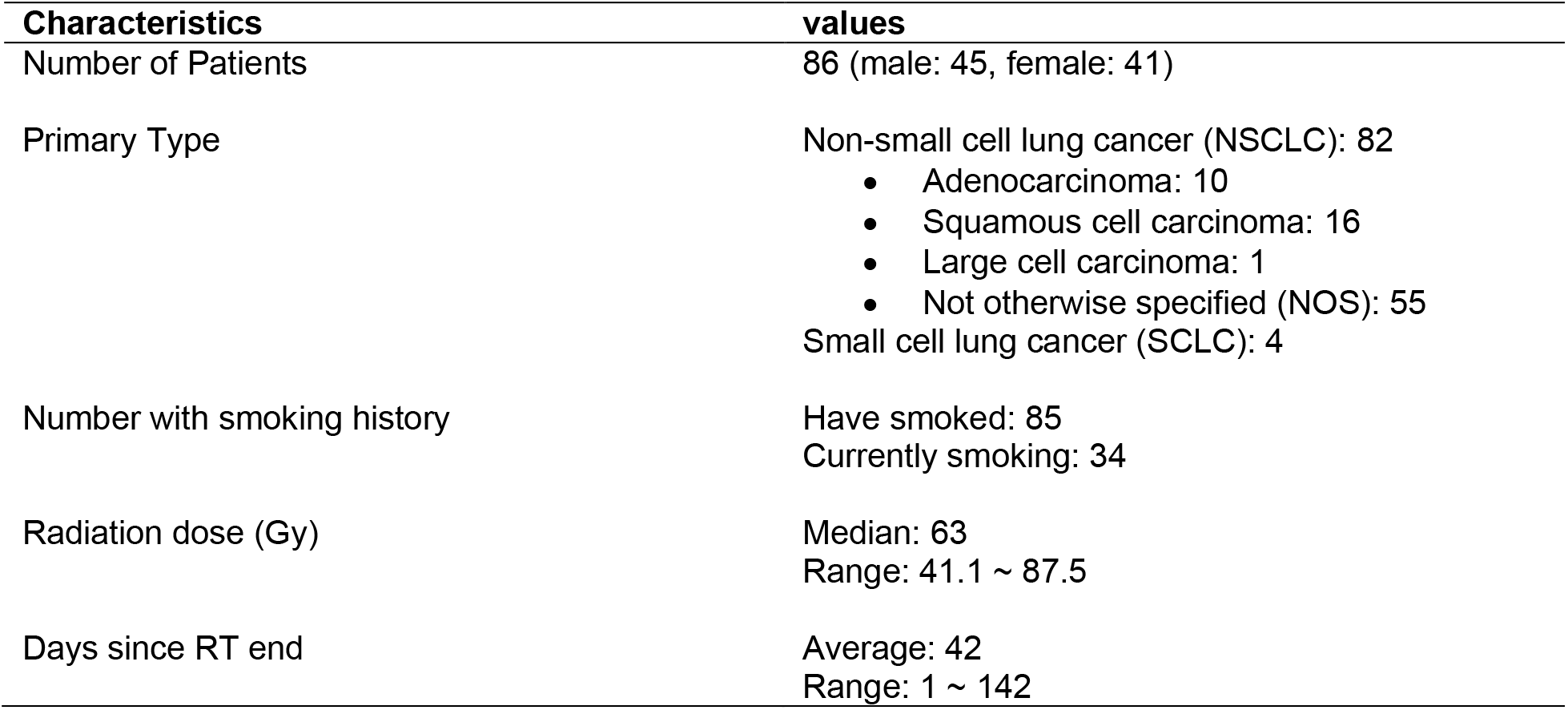
Patient characteristics and radiation dose

### Grading system

In this study, we used a grading system that was developed to assess the severity of RILD on each CT scan based on the systems proposed by Libshitz and Shuman^14^ and Bush et al^18,19^. Grading (0 to 5) was assigned as follows:

- Grade 0 – lung appearance similar to the baseline CT scan (pre-treatment scan)
- Grade 1 – ground glass opacities involving the irradiated portions of the lung without consolidation
- Grade 2 – patchy consolidation that is contained, but does not conform to the shape of the radiation portal
- Grade 3 – discrete consolidation that conforms to the shape of the portal and outlines the portal, but does not uniformly involve it
- Grade 4 – solid consolidation that conforms to and totally involves the radiated portion of the lung
- Grade 5 – solid consolidation that involves the radiated portion of the lung and is associated with volume loss and traction bronchiectasis, CT signs of lung fibrosis.

Radiologists (MB and EGO) assigned all scans to one of these six categories.

### Feature extraction

First, the delineation of the whole lung was performed automatically based on the intensity threshold (Hounsfield units < -200) of the CT image (Fig. 1b). We applied the median filter over the delineated whole-lung image. The median filter is a nonlinear filtering technique to remove noise from an image. The window size of this filter was 13 × 13, and sliding it over the lung image allowed us to reduce the signal of the noise or small vessels in the background of the lung. Then, the abnormal area, or region of interest (ROI), inside the lung on the CT image was delineated semi-automatically by setting a threshold. The median filter was used only for delineating an ROI. The radiomic features were extracted from CT images of patients before and after RT. Fig. 1 shows an example of a lung CT image and its median filtered map. We used a low-pass filter called a maximally flat magnitude filter (Butterworth filter) for smoothing images as a preprocessing step. Low pass filtering has been shown to help mitigate the impact of differing pixel sizes in the original image data sets.^20^ This filter removes noise from the image and has the added benefit of reduced ringing and a gradual attenuation of higher frequencies^21-23^.

**Fig. 1.**
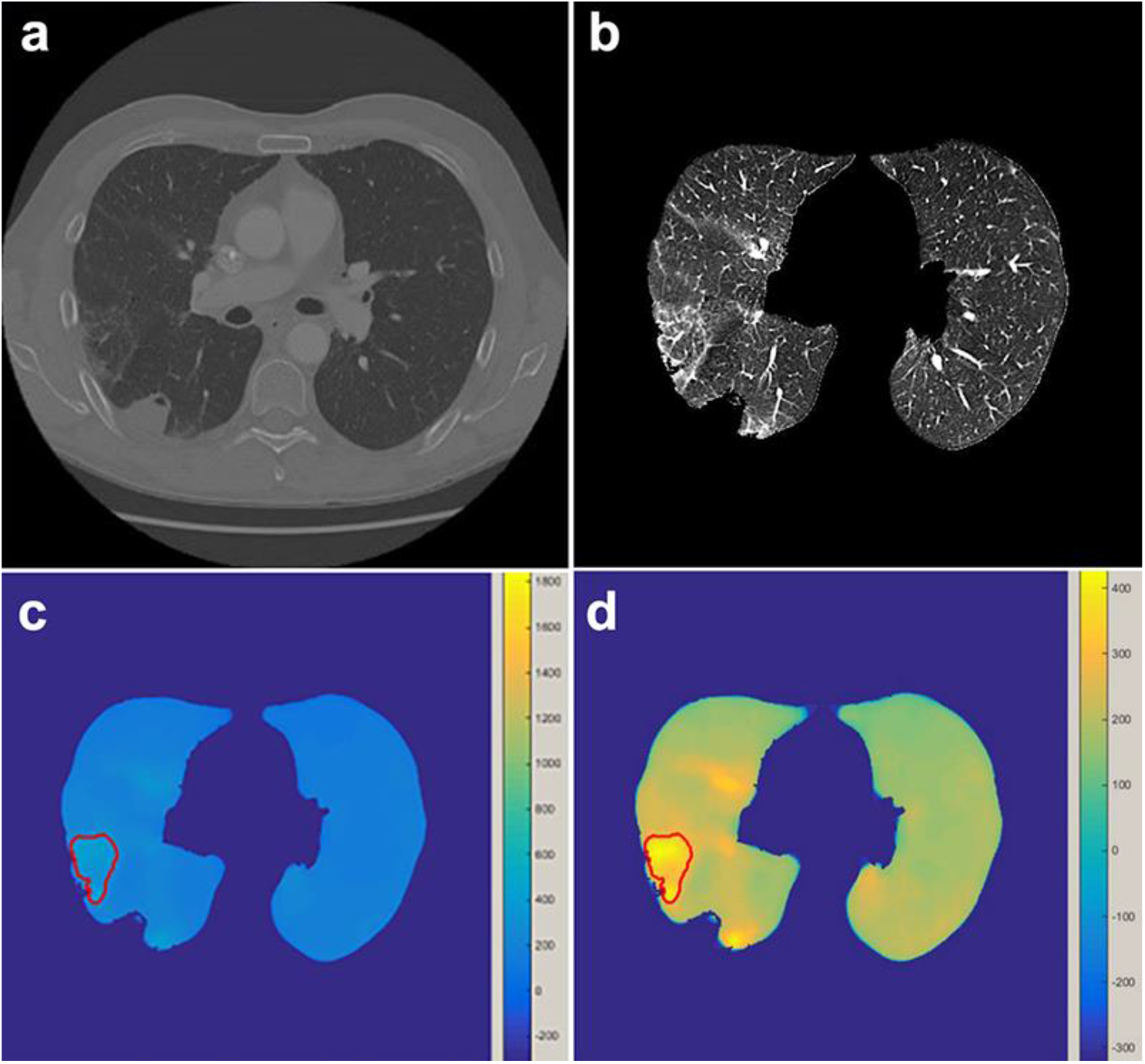
Examples of (a) CT image that has ground glass opacities and (b) lung image delineated with a threshold. (c) A median filtered heat map was generated with a median filter that slides over the lung image. Fig (d) shows a rescaled heat map.

We extracted a total of 138 quantitative image features from the delineated ROIs using our in-house imaging software called IBEX ^24^. This software was designed based on MATLAB (v8.1.0, MathWorks, Natick, MA) and is available at http://bit.ly/IBEX_MDAnderson.

The image features consisted of 55 intensity direct features, 66 texture features, and 17 shape features. The detailed features are listed in Table 2. For texture features, we used a gray-level co-occurrence matrix (GLCM) that measures the spatial relationship of pixels and enables characterization of the texture of an image with various statistics such as energy, contrast, and correlation. For example, energy measures the homogeneity of an image by summing squared elements of the GLCM. A more homogeneous image has fewer gray levels with higher pixel elements of the GLCM. Contrast measures the luminance (differences in gray-level intensity values) present in an image. Correlation of the GLCM measures the gray-level linear dependence of pixels at specified positions. We calculated GLCM at 0, 45, 90, and 135 degree and used the average of these four matrices with three pixel offsets: 1, 4, and 7 pixels^25,26^.

**TABLE 2.**
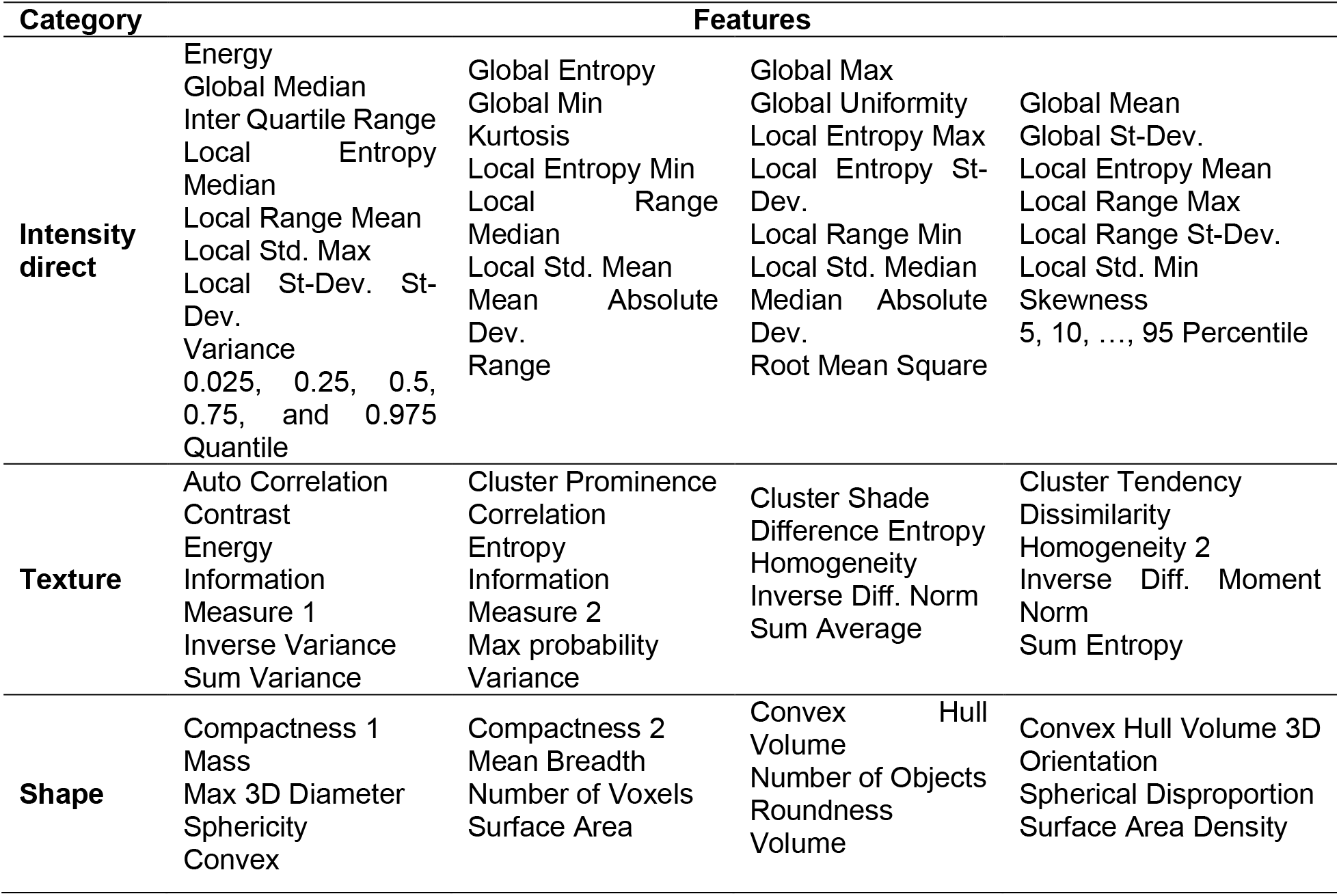
Radiomic features extracted from the delineated regions of interest

### Data analysis

In this study, we compared lung damage of six different grades (0 ∼ 5) in three steps. First, we compared scans from patients with grade 0 lung damage to those with grade 1 ∼ 5 lung damage. In this step (step 1), we used delta radiomic features^16,23^ (Eq.[1]), which measured the difference between pre-treatment and post-treatment scans in features.

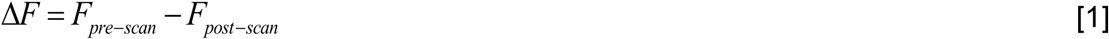

where *F*_*pre-scan*_ and *F*_*post-scan*_ are features extracted from the pre-treatment scan and post-treatment scan, respectively.

With these delta features, the delta feature values for grade 0 scans (similar to pre-treatment scan) were minimized and could be differentiated from the feature values for other grades. For step one, forty patient scans were used for comparison, with 20 scans consisting of patients with grade 0 lung damage and 20 scans with grade 1 ∼ 5 lung damage were used for comparison. Next (step 2), we compared patients with grade 1 lung damage to those with grade 2 ∼ 5 lung damage. Twenty-five scans of patients with grade 1 lung damage and 25 scans of patients with grade 2 ∼ 5 lung damage were used in this step. Last (step 3), we compared patients with grade 2∼4 lung damage (20 scans) to those with grade 5 lung damage (20 scans). Fig. 2 shows the flow chart for each step.

**Fig. 2.**
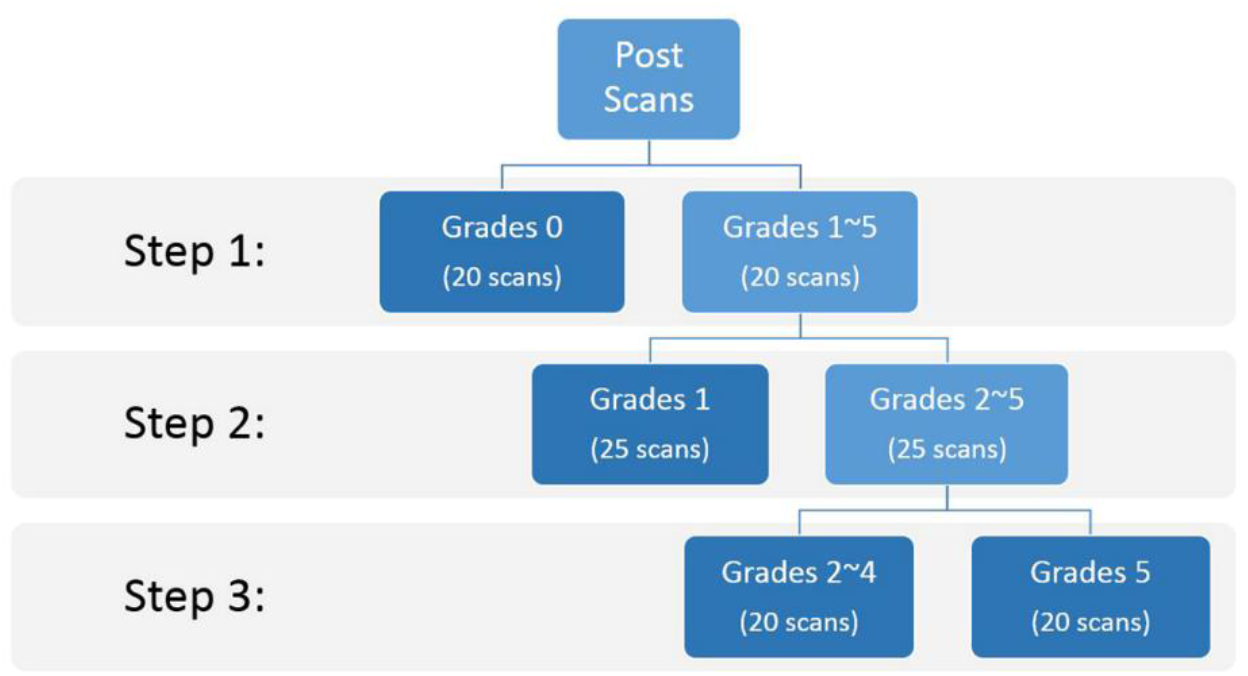
Flowchart illustrating the sequential comparisons of different grades of lung damage

We also validated the model using a separate dataset that had not been used for training and testing. The validation dataset consisted of ten scans of grade 0 and ten scans of grades 1∼5 for step 1, ten scans of grade 1 and ten scans of grades 2∼5 for step 2, and ten scans of grades 2∼4 and ten scans of grade 5 for step 3. In this study, we used a random forest algorithm as a classifier, which is an ensemble decision-trees learning method that yields a prediction value for classification^27^. All statistical analyses were computed in R (R Core Team, Vienna, Austria, https://www.R-project.org/).

## RESULTS

### Step 1: Grade 0 vs Grades 1∼5

In this step, we randomly selected 20 pre-treatment and post-treatment CT scans, respectively. The classification accuracy was 70%, and the class error rates for pre-treatment and post-treatment were 30% and 30%, respectively. Table 3 shows the confusion matrix for step 1.

**TABLE 3.** Confusion matrix for comparison of pre- and post-treatment scans (step 1).

|  | Predicted:<br>0 | Predicted:<br>1 | Class error |
| --- | --- | --- | --- |
| Actual: 0 | 14 | 6 | 0.30 |
| Actual: 1 | 6 | 14 | 0.30 |

0: pre-treatment, 1: post-treatment

### Step 2: Grade 1 vs Grades 2∼5

In this step, 25 scans from patients with grade 1 lung damage and 25 scans of grade 2 ∼ 5 lung damage were compared. The classification accuracy was 90%, and the class error rates for grade 1 and grades 2∼5 were 8% and 12%, respectively. Table 4 shows the confusion matrix for step 2.

**Table 4.**
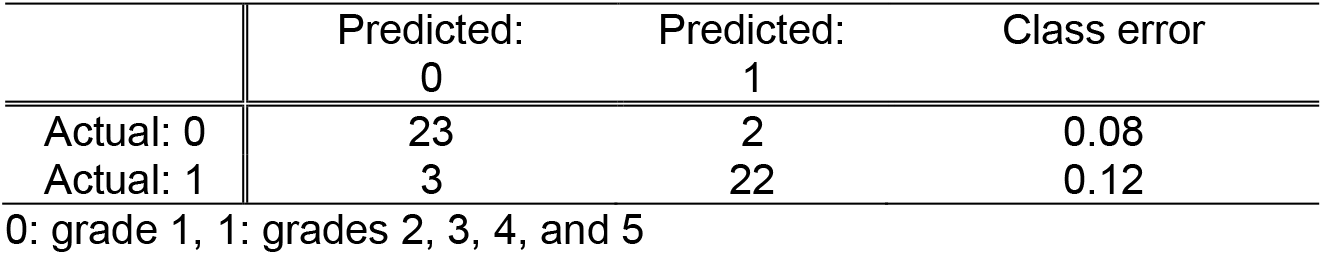
Confusion matrix for comparison between grade 1 and grades 2 ∼ 5 (step 2)

### Step 3: Grades 2 ∼ 4 vs Grade 5

In the last step, 20 scans from patients with grade 2∼4 lung damage and 20 scans of grade 5 lung damage were compared. The classification accuracy was 80%, and the class error rates for grades 2∼4 and grade 5 were 15% and 25%, respectively. Table 5 shows the confusion matrix for step 3.

**Table 5.** Confusion matrix for comparison between grades 2∼4 and grade 5 (step 3)

|  | Predicted:<br>0 | Predicted:<br>1 | Class error |
| --- | --- | --- | --- |
| Actual: 0 | 17 | 3 | 0.15 |
| Actual: 1 | 5 | 15 | 0.25 |

0: grades 2~4, 1: grade 5

### Validation

A total of 60 scans in 24 patients were used for the validation test. The classification accuracies for the validation dataset were 15%, 5%, and 15% for steps 1, 2, and 3, respectively. Table 6 shows the final classification results for the validation dataset.

**TABLE 6.** Validation results. Twenty scans were used for each step (step1: grade 0 vs grade 1∼5, step2: grade 1 vs grade 2∼5, and step3: grade 5 vs grade 2∼4)

| Scan Index | Actual Label | Step 1 Prediction | Step 2 Prediction | Step 3 Prediction |
| --- | --- | --- | --- | --- |
| 1 | 0 | 0 | 0 | 0 |
| 2 | 0 | 0 | 0 | 0 |
| 3 | 0 | 0 | 0 | 1 |
| 4 | 0 | 0 | 0 | 0 |
| 5 | 0 | 1 | 0 | 0 |
| 6 | 0 | 0 | 0 | 0 |
| 7 | 0 | 0 | 1 | 1 |
| 8 | 0 | 1 | 0 | 1 |
| 9 | 0 | 0 | 0 | 0 |
| 10 | 0 | 0 | 1 | 0 |
| 11 | 1 | 0 | 1 | 1 |
| 12 | 1 | 1 | 1 | 1 |
| 13 | 1 | 1 | 1 | 1 |
| 14 | 1 | 1 | 1 | 1 |
| 15 | 1 | 1 | 1 | 1 |
| 16 | 1 | 1 | 1 | 1 |
| 17 | 1 | 1 | 1 | 1 |
| 18 | 1 | 1 | 1 | 1 |
| 19 | 1 | 1 | 1 | 1 |
| 20 | 1 | 1 | 1 | 1 |
| Error rate |  | 15% | 5% | 15% |

Label 0 is grade 0 for step 1, grade 1 for step 2, and grades 2~4 for step 3. Label 1 is grades 1~5 for step 1, grades 2~5 for step 2, and grade 5 for step 3.

## Discussion

This study demonstrated that the radiomics features measured from lung damage could be used for identifying the severity of lung damage after chemoradiotherapy. Chemotherapy and radiotherapy are commonly used to treat lung cancer but may result in lung damage. RILD can have a variety of appearances, especially depending on the severity of the lung damage. In this study, we used a more specific grading system (consisting of 6 categories) than the grading system in which the damage is divided into two stages such as early inflammatory (radiation pneumonitis) and later complications of chronic scarring (radiation fibrosis).

In the present study, we performed the classification of the severity of the lung damage in three steps. Grade 0 is defined as the lung in the post-treatment CT scan appearing similar to the lung in the pre-treatment CT scan. This indicates that the features from scans with grade 0 will have between patient variability depending on their pre-treatment scan. In the first step, we used delta features to remove grade 0 variability between patients. Since the other grades for RILD have specific shape and texture criteria that are irrespective of the patient, we do not need to calculate delta features for the subsequent data analysis steps. The classification between grade 0 and grades 1 to 5 was performed and gave us a classification accuracy of 70% for the test dataset and 85% for the validation dataset. These classification accuracies are relatively low compared to the classification accuracies of steps 2 and 3. This might be because grade 0 was assigned to scans in which the post-treatment appearance of the lung was similar to that of the baseline CT scan; thus, grade 0 could look different for each patient. Although we used delta features to remove grade 0 variability between patients, this variability may not completely removed.

In step 2, the objective was to classify between patients with grade 1 and grade 2∼5 lung damage. The classification accuracies for this step were 90% for the test dataset and 95% for the validation dataset. This level of accuracy was the highest among all of the steps. In this step, ground-glass opacity (grade 1) was compared with discrete or solid consolidation (grade 2∼5). According to these results, the features from grade 1 patents (ground-glass opacity) will be the most distinctive from the features from other grades. In this step, the abnormal area was detected and delineated semi-automatically using a median filter, which is a digital filtering technique often used to remove noise from an image. This median filter worked very well for the detection of abnormal areas, especially ground-glass opacities (grade 1) and gave us consistent procedure for delineating an ROI. This automatic detection of abnormal areas could have potential benefits in clinical practice as it can save time and improve contouring consistency. Furthermore, early detection and treatment of ground-glass opacity may improve the prognosis of lung cancer. For step 3, the patients with grade 2∼4 and grade 5 lung damage were classified, with classification accuracies of 80% for the test dataset and 85% for the validation dataset.

In this study, we examined the feasibility of radiomics with a machine-learning algorithm for identifying the severity of RILD. Some limitations must be noted. First, we contoured an ROI semi-automatically using a median filter that detects ground-glass opacities and abnormalities inside the lung very well (Fig. 1). However, if solid consolidation presents on the lung wall, then initial contouring of the lung with intensity threshold will exclude this solid consolidation, and the ROI has to be contoured manually. Since this process is not fully automatic, it is time-consuming to contour all ROIs. Furthermore, due to their indistinct boundaries, manual detection and segmentation of RILD is problematic. Additionally, we used two different slice thicknesses (2.5mm and 3.8mm). Although we tried to use similar slice thicknesses, it might affect the quantification of CT image features ^28^. Another limitation for this study is that this is the retrospective study and the data were collected between 2001 and 2012. This indicated that there might be the possible impact of inter- and intra-scanner variabilities on the radiomics features ^20,29^.

Our study shows that radiomics could be a promising method for identifying CT textural changes for RILD, and it could be a potential tool for predicting the severity of RILD. For future study, radiomics with an advanced machine-learning algorithm and fully automatic contouring method could increase the classification accuracy. Such a setup will help diagnostic radiologists identify and differentiate abnormalities from RILD.

## CONCLUSION

We investigated the feasibility of radiomics for identifying the severity of lung damage after chemoradiotherapy that could reflect for radiation-induced lung damage (RILD). Our preliminary study shows that the classification accuracy for classifying the grade of RILD is greater than 70%, and the accuracy is greater than 90% for identifying ground-glass opacity (grade 1) lung damage over other grades. This preliminary study using a radiomics approach supports potential tools for identifying the severity of RILD on CT images that will help diagnostic radiologists identify and classify abnormalities from RILD.

## Data Availability

All data produced in the present study are available upon reasonable request to the authors.

## ACKNOWLEDGEMENTS

The authors would like to thank Sunita Patterson of MD Anderson’s Department of Scientific Publications for scientific editing. The funding for this work was provided by the generous support from the Scurlock Foundation to the Center for Radiation Oncology Research at the University of Texas MD Anderson Cancer Center.

## CONFLICT OF INTEREST DISCLOSURE

The authors have no COI to report.

## Data Availability

The datasets generated during and/or analyzed during the current study are available from the corresponding author on reasonable request.

## Author Contributions

Project conception and design were by J.L., L.C., S.L. The data collection and preprocessing were performed by J.L., M.B., E.G.O., and S.L. The software programming, statistical analysis, and interpretation were performed by J.L. The manuscript was written by J.L. and all authors reviewed the manuscript.

## Notes

### Competing Interest Statement

The authors have declared no competing interest.

### Funding Statement

This study did not receive any funding

### Author Declarations

This study was approved by the Institutional Review Boards of the University of Texas MD Anderson Cancer Center.

